# Benchmarking COVID-19 Mortality in the United States

**DOI:** 10.1101/2020.09.30.20204586

**Authors:** Ruth Etzioni, Elan Markowitz, Ivor S. Douglas

## Abstract

On September 22^nd^ the US officially recorded 200,000 COVID-19 deaths. It is unclear how many deaths might have been expected in the case of an early and effective response to the pandemic. We aim to provide a best-case estimate of COVID-19 deaths in the US by September 22^nd^ using the experience of Germany as a benchmark. Our methods accommodate the differences in demographics between Germany and the US. We match cumulative incidence of COVID-19 deaths by age group in Germany to non-Hispanic whites in the US and project the implied number of deaths in this population and among the black and Hispanic populations under observed racial/ethnic disparities in cumulative COVID-19 mortality in the US. We estimate that if the US had been as successful as Germany in managing the pandemic we would have expected 22% of the deaths actually recorded. The number of deaths would have been lower by a further one-third if we could have eliminated racial/ethnic disparites in COVID-19 outcomes. We conclude that almost 80 percent of the COVID-19 deaths in the US by September 22^nd^ could have been avoided with an early and effective response producing similar age-specific death rates among non-Hispanic whites as in Germany.

## Introduction

As the US surpasses 200,000 officially recorded COVID-19 deaths, there is a sense that mortality could have been markedly reduced had we acted earlier and more effectively. An early modeling study^1^ concluded that if a national shutdown had occurred just one week earlier, 36,000 lives could have been saved by early May. At this point, however, we find ourselves lacking a benchmark against which to compare the current number of lives lost. How many deaths could we have averted by now had the response been optimal?

This article aims to provide a reasonable estimate of the number of COVID-19 deaths expected in a best-case scenario – had the US responded with widely accessible rapid turn-around testing, effective contact tracing and isolation of exposed individuals, and compliance with social distancing and masking. We take as our model the country of Germany whose initial response to the virus has been credited with producing a notably low death rate^2^. Essentially, we address the following question: If the US had been as successful as Germany in managing the pandemic, how many deaths would we have expected to date? By “as successful as Germany,” we mean: “if the age-specific cumulative incidence of COVID-19 deaths as a fraction of the population in the US were the same as in Germany.”

It is not enough to simply translate the COVID-19 death rate per capita in Germany into a number of US deaths, because the age and demographic structures of the two countries are quite different. The vast majority of Germans are European, fewer than 1% are African, and 5% are Middle Eastern or North African^3^. In the US, 18% are Hispanic or Latino and 13% are African American^4^. The German population skews older^3^ with 22% over 65 (versus 16% in the US). Our methods accommodate demographic differences between the two populations and project expected lives lost based on the German experience while preserving the racial/ethnic disparities in COVID-19 outcomes observed in this country.

## Methods

We obtained cumulative incidence of German COVID-19 deaths by age through September 22 from the Robert Koch Institute^5^. We then matched the age-specific cumulative incidence of COVID-19 deaths in Germany^5^ to US non-Hispanic whites by age group (Figure 1). African-American and Hispanic/Latino populations in the US have had markedly higher COVID-19 mortality than non-Hispanic whites.^6^ For these minority populations we offer two projections. First, we use the German death rates inflated by the corresponding death rate ratios (relative to non-Hispanic whites) by age which we calculate using publicly available CDC data on age-specific death counts relative to population by race/ethnicity for the US overall.^7^ As a counterpoint, we use the German death rates without inflation for these minority populations. This analysis reflects a counterfactual in which factors driving disparities are fully addressed. In both cases, we assume other race/ethnicities have mortality that is similar to non-Hispanic whites. We then estimate a weighted average of the expected death rates by age and race/ethnicity using US census weights^8^. Data and calculations are provided in the Supplemental Table.

**Figure 1:**
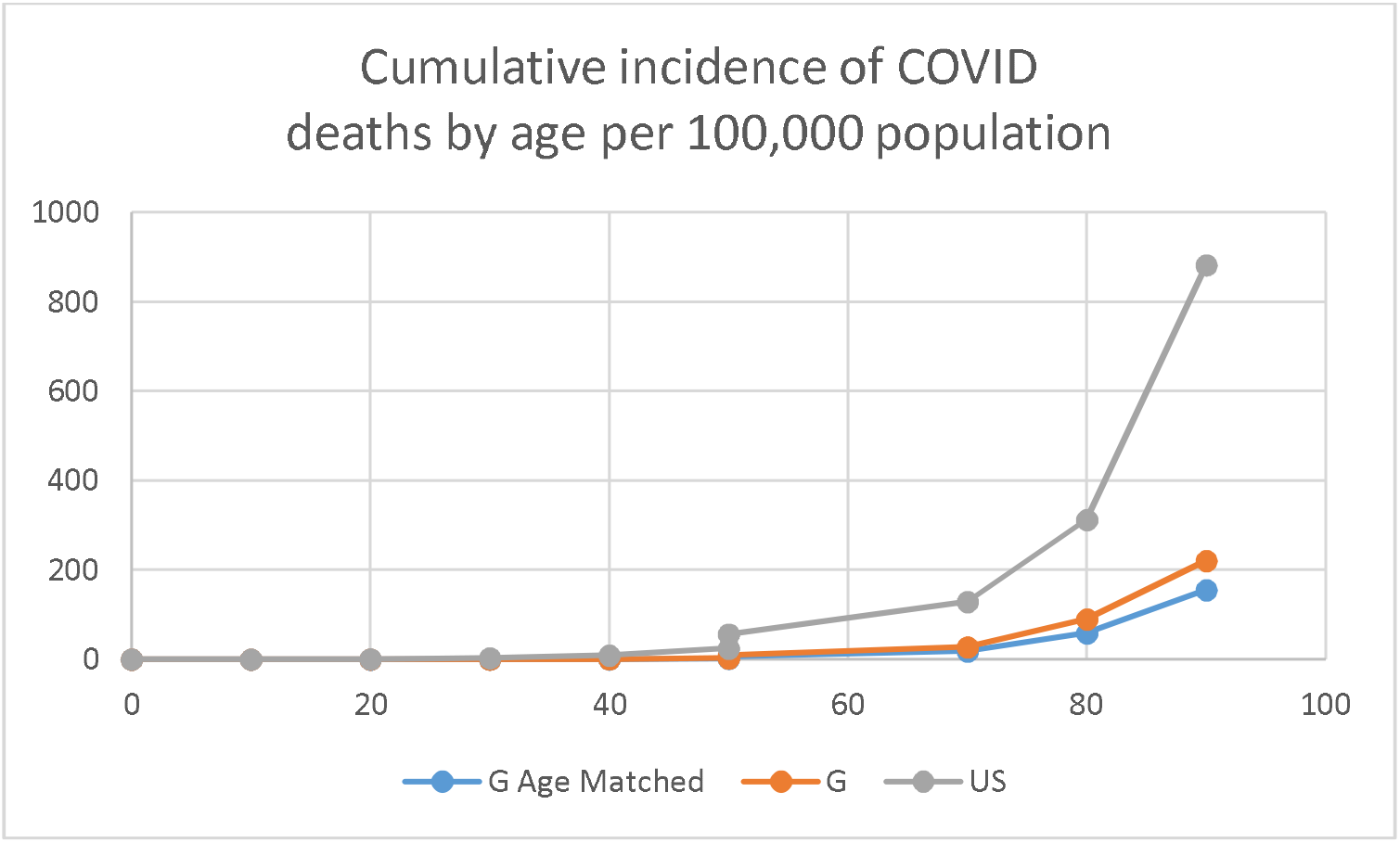
Cumulative incidence of COVID-19 deaths in Germany and the US by age. German death rates are provided by age decade (0-9, 10-19….,90+). The figure shows observed German death rates (G) plotted against the lower limit of each age group and US cumulative death rate plotted against the midpoint of each US age group as well as an age-matched version (G age matched) of the German death rates.

Matching German age-specific cumulative death rates through September 22 to US age groups, and preserving the aforementioned disparities, we estimate that over the same time period the US would have expected to 43,187 recorded deaths rather than the recorded cumulative total of 200,000 by this date. If we could have eliminated racial and ethnic disparities so that death rates for black and Hispanic populations matched those in non-Hispanic whites, this number would be 28,835. Obviously, this latter estimate is unrealistically low, but we calculate it to make the point that the success of a country in managing a pandemic lies not only in its absolute burden of deaths, but also in its efforts to minimize health inequities.

## Discussion

Naturally, there are numerous caveats in interpretation of these findings. First and most important, the analysis uses age specific death rates from Germany to represent mortality expected in the US if the US could have responded in a manner that produced mortality patterns similar to those observed in Germany. While age and race/ethnicity are primary drivers of mortality risk, there are many relevant differences between the US and Germany not captured here. These include differences in population density and travel patterns as well as culture, that could impact compliance with mitigation measures. Also of interest due to its prognostic significance, is the frequency of obesity – just under one fourth of German adults are obese^10^, whereas among US adults, the fraction obese is around 40 percent^11^. These differences suggest that our results may reflect an optimistic version of what we would expect even if the US had acted similarly in terms of mitigation policies and resources. Given these caveats, our analysis could be considered a thought experiment designed to provide a first quantification of a best-case scenario in this country.

A further key point concerns what is meant by a COVID-19 death. Whether a patient is determined to have died of the virus is not a simple yes-no decision^12^. The definition of a COVID-19 death is not the same across countries, and even in the US varies across states and (in some cases) changes over time^13^. If the definition could be standardized the results might differ from those presented here.

A legitimate question concerns our selection of Germany as a model. Other countries such as South Korea or New Zealand could have been used, but we selected Germany because of its demographic and economic similarities with the US and its record of early action on COVID-19. Notably, the first German COVID-19 case was diagnosed on January 27, less than one week after the first US case was identified on January 21.

While we accounted for elevated mortality in the largest minority racial/ethnic groups, we did not model relative death rates in other smaller groups. Our assumption that overall these have similar experience to non-Hispanic whites may be optimistic; for example, American Indian and Alaska Native persons have a markedly higher incidence^14^ than non-Hispanic whites. However, they represent a small minority of the remaining population, which is mostly Asian with mortality similar to non-Hispanic whites. Consequently, we expect that accounting for smaller racial/ethnic groups will affect results only modestly.

We conclude that in the presence of a highly effective response to the COVID-19 pandemic, the lives lost by September 22 could have been as low as one fifth of the total recorded deaths in the US. Going forward, this result will change with changes in transmission, behavior and policy in both countries. We are not doomed to perpetuate our divergent histories; we can still narrow the gap as we navigate how to coexist with COVID-19.

Observed German death rates (to September 22) are plotted against the midpoint of the closest US age group (e.g. German death rate for 60-69 at age 60). To match German death rates to US age groupings, we use the following formula: rate(i)=(rate(j)+rate(k))/2 where i reflects a US age group e.g. [5-14], and j and k are the encompassing German age groups e.g. [0-9] and [10-19]. For ages 85+ we apply this formula using j=[80-89] and k=90+ and for ages [0-4] we use the German [0-9] death rate. US death rates are based on CDC data through September 19^15^.

## Data Availability

All data is publicly available. German death rates are from the Robert Koch Institute. US population and deaths by race/ethnicity are from the CDC.

https://www.rki.de/EN/Content/infections/epidemiology/outbreaks/COVID-19/COVID19.html

https://data.cdc.gov/NCHS/Deaths-involving-coronavirus-disease-2019-COVID-19/ks3g-spdg

## Notes

### Competing Interest Statement

The authors have declared no competing interest.

### Funding Statement

This project was not funded by any specific mechanism

## References

1. Pei S, Kandula S, Shaman J. Differential Effects of Intervention Timing on COVID-19 Spread in the United States. medRxiv. May 2020. doi:10.1101/2020.05.15.20103655

2. A German Exception? Why the Country’s Coronavirus Death Rate Is Low - The New York Times. https://www.nytimes.com/2020/04/04/world/europe/germany-coronavirus-death-rate.html. Accessed September 24, 2020.

3. Wikipedia, Wikimedia Foundation. Demographics of Germany. https://en.wikipedia.org/wiki/Demographics_of_Germany. Accessed September 23, 2020.

4. U.S. Census Bureau QuickFacts: United States. https://www.census.gov/quickfacts/fact/table/US/PST045219. Accessed September 24, 2020.

5. Robert Koch Institut - COVID-19. https://www.rki.de/EN/Content/infections/epidemiology/outbreaks/COVID-19/COVID19.html. Accessed September 29, 2020.

6. Race gaps in COVID-19 deaths are even bigger than they appear. https://www.brookings.edu/blog/up-front/2020/06/16/race-gaps-in-covid-19-deaths-are-even-bigger-than-they-appear/. Accessed September 24, 2020.

7. Deaths involving coronavirus disease 2019 (COVID-19) by race and Hispanic origin group and age, by state | Data | Centers for Disease Control and Prevention. https://data.cdc.gov/NCHS/Deaths-involving-coronavirus-disease-2019-COVID-19/ks3g-spdg. Accessed September 15, 2020.

8. 2019 Population Estimates by Age, Sex, Race and Hispanic Origin. https://www.census.gov/newsroom/press-kits/2020/population-estimates-detailed.html. Accessed September 24, 2020.

9. US Historical Data | The COVID Tracking Project. https://covidtracking.com/data/national. Accessed September 29, 2020.

10. OECD. The Heavy Burden of ObesityThe Economics of Prevention. https://www.oecd.org/germany/Heavy-burden-of-obesity-Media-country-note-GERMANY.pdf. Accessed September 24, 2020.

11. CDC. Adult Obesity Facts. https://www.cdc.gov/obesity/data/adult.html. Accessed September 24, 2020.

12. How COVID-19 Deaths Are Counted - Scientific American. https://www.scientificamerican.com/article/how-covid-19-deaths-are-counted1/. Accessed September 24, 2020.

13. Seaman J, Burness A. Colorado changes how coronavirus deaths are reported as COVID-19 fatalities become political flashpoint. The Denver Post. May 15, 2020.

14. Hatcher SM, Agnew-Brune C, Anderson M, et al. COVID-19 Among American Indian and Alaska Native Persons - 23 States, January 31-July 3, 2020. MMWR Morb Mortal Wkly Rep. 2020;69(34):1166–1169. doi:10.15585/mmwr.mm6934e1

15. CDC. US Mortality by Age. https://data.cdc.gov/NCHS/Provisional-COVID-19-Death-Counts-by-Sex-Age-and-S/9bhg-hcku. Accessed September 29,2020

