## Supplemental Table for "Benchmarking COVID-19 Mortality in the United States"

Calculations for "Benchmarking COVID-19 Mortality in the United States" by Etzioni R, Markowitz E, and Douglas IS.

Projected Deaths under German response. Assumes German death rate represents NHW population. German death rate is using mean of neighboring age groups.

| | | Germany | | | | United States | | | | |
|---|---|---|---|---|---|---|---|---|---|---|
| Age Group | Population | Deaths | Death Rate (per person) | Age Group | Population | Deaths | Death Rate (per person) | German Age-Matched Death Rate | If German Death Rate | If Age-Matched German Death Rate |
| 0-9 | 7588635 | 1 | 1.31776E-07 | 0-4 | 19576683 | 35 | 1.78784E-06 | 1.31776E-07 | 2.579737067 | 2.57974 |
| 10-19 | 7705657 | 1 | 1.29775E-07 | 5-14 | 40994163 | 31 | 7.56205E-07 | 1.30775E-07 | 5.320008794 | 5.36103 |
| 20-29 | 9800607 | 10 | 1.02034E-06 | 15-24 | 42687510 | 353 | 8.2694E-06 | 5.7506E-07 | 43.55598587 | 24.54787 |
| 30-39 | 10646445 | 26 | 2.44213E-06 | 25-34 | 45940321 | 1457 | 3.17151E-05 | 1.73124E-06 | 112.1922244 | 79.53360 |
| 40-49 | 10426257 | 82 | 7.86476E-06 | 35-44 | 41659144 | 3809 | 9.14325E-05 | 5.15344E-06 | 327.6391334 | 214.68808 |
| 50-59 | 13474166 | 347 | 2.5753E-05 | 45-54 | 40874902 | 10057 | 0.000246043 | 1.68089E-05 | 1052.650754 | 687.06101 |
| 60-69 | 10302411 | 914 | 8.87171E-05 | 55-64 | 42448537 | 23991 | 0.000565178 | 5.7235E-05 | 3765.910991 | 2,429.54377 |
| 70-79 | 7685929 | 2120 | 0.000275829 | 65-74 | 31483433 | 40613 | 0.00128998 | 0.000182273 | 8684.035197 | 5,738.57699 |
| 80-89 | 4594163 | 4137 | 0.00090049 | 75-84 | 15969872 | 49871 | 0.003122818 | 0.00058816 | 14380.71755 | 9,392.83349 |
| 90+ | 794943 | 1754 | 0.002206448 | 85+ | 6604958 | 58253 | 0.008819587 | 0.001553469 | 14573.49311 | 10,260.59742 |
| | | 9392 | | | 328239523 | 188470 | | | **42,948** | **28,835** |
| | | (9/22/20) | | | | (9/19/20) | | | | |
| | | RKI numbers | | | | CDC numbers | | | | |

| Age | G Age Matched | G | US |
|---|---|---|---|
| 0 | 0.0131776 | 0.0131776 | 0.17878412 |
| 10 | 0.01307754 | 0.012977479 | 0.07562052 |
| 20 | 0.057505988 | 0.102034496 | 0.82693978 |
| 30 | 0.173123735 | 0.244212974 | 3.17150592 |
| 40 | 0.515344444 | 0.786475914 | 9.14325076 |
| 50 | 1.680887226 | 2.575298538 | 24.6043403 |
| 50 | 5.723504138 | 8.871709739 | 56.5178489 |
| 70 | 18.22729114 | 27.58287255 | 128.998003 |
| 80 | 58.81595978 | 90.04904702 | 312.281777 |
| 90 | 155.3468988 | 220.6447506 | 881.958674 |

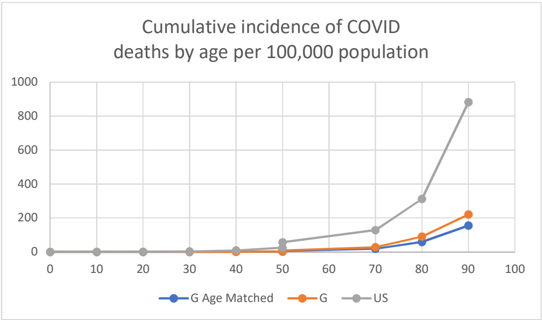


Sources:

German mor Robert Koch Institut - COVID-19. https://www.rki.de/EN/Content/infections/epidemiology/outbreaks/COVID-19/COVID19.html. Accessed September 29, Data current as of September 22.

US mortality US Mortality by Age (https://data.cdc.gov/NCHS/Provisional-COVID-19-Death-Counts-by-Sex-Age-and-S/9bhg-hcku) accessed 9/29

Calculations for "Benchmarking COVID-19 Mortality in the United States" by Etzioni R, Markowitz E, and Douglas IS.

Projected Deaths under German response. Separated By Age and Race (Assumes German death rate represents White population and US relative rates between races are unchanged). German death rate is using mean of neighboring age groups. Relative race dispari

| Age Group | White Pop | White Deaths | White Death Rate | Hispanic Pop | Hispanic Deaths | Hispanic Death Rate | Hispanic/White Death Rate rat | Black Pop | Black Deaths | Black Death Rate | Black/White Death Rate ratio |
|---|---|---|---|---|---|---|---|---|---|---|---|
| 0-4 | 10,566,427 | 9 | 8.51754E-07 | 4571556 | 14 | 3.06241E-06 | 3.595420076 | 3594764 | 5 | 1.39091E-06 | 1.6329966 |
| 5-14 | 22,144,655 | 3 | 1.35473E-07 | 9506446 | 14 | 1.47268E-06 | 10.87070008 | 7409939 | 10 | 1.34954E-06 | 9.9616902 |
| 15-24 | 23,910,455 | 58 | 2.42572E-06 | 8934485 | 139 | 1.55577E-05 | 6.413648034 | 7340656 | 92 | 1.25329E-05 | 5.1666947 |
| 25-34 | 26,050,842 | 217 | 8.32987E-06 | 8582496 | 585 | 6.8162E-05 | 8.182844295 | 7566166 | 397 | 5.24704E-05 | 6.2990734 |
| 35-44 | 24,146,655 | 510 | 2.11209E-05 | 7836043 | 1747 | 0.000222944 | 10.5555993 | 6068006 | 1002 | 0.000165128 | 7.8182314 |
| 45-54 | 25,583,095 | 1894 | 7.40333E-05 | 6508843 | 3994 | 0.000613627 | 8.288527321 | 5623226 | 2673 | 0.00047535 | 6.4207614 |
| 55-64 | 29,856,071 | 6920 | 0.000231779 | 4642018 | 7085 | 0.001526276 | 6.585057853 | 5323702 | 6498 | 0.001220579 | 5.2661416 |
| 65-74 | 23,789,043 | 16088 | 0.000676278 | 2601307 | 8768 | 0.003370613 | 4.984066695 | 3300966 | 10038 | 0.003040928 | 4.4965668 |
| 75-84 | 12,467,083 | 26104 | 0.002093834 | 1217089 | 7993 | 0.006567309 | 3.136499811 | 1451540 | 9610 | 0.006620555 | 3.1619294 |
| 85+ | 5,253,844 | 38495 | 0.007327016 | 478035 | 6205 | 0.012980221 | 1.771556223 | 542174 | 7329 | 0.013517801 | 1.8449257 |
| | 203,768,170 | | | 54,878,318 | | | | 48,221,139 | | | |

Germany / German Projected to US (extrapolate white pop to unnaccounted for)

| Age Group | White death rate (to | Black Est. | Hisp Est | White Deaths | Black Deaths | Hispanic Deaths | |
|---|---|---|---|---|---|---|---|
| 0-4 | 1.31776E-07 | 2.1519E-07 | 4.7379E-07 | 1.392401532 | 0.7735564 | 2.165957939 | |
| 5-14 | 1.30775E-07 | 1.3027E-06 | 1.4216E-06 | 2.895976027 | 9.6532534 | 13.51455479 | |
| 15-24 | 5.7506E-07 | 2.9712E-06 | 3.6882E-06 | 13.74994329 | 21.810255 | 32.95245029 | |
| 25-34 | 1.73124E-06 | 1.0905E-05 | 1.4166E-05 | 45.10019078 | 82.510487 | 121.5834636 | |
| 35-44 | 5.15344E-06 | 4.0291E-05 | 5.4398E-05 | 124.438445 | 244.48494 | 426.2626732 | |
| 45-54 | 1.68089E-05 | 0.00010793 | 0.00013932 | 430.0229758 | 606.89093 | 906.817194 | |
| 55-64 | 5.7235E-05 | 0.00030141 | 0.0003769 | 1708.813459 | 1604.6055 | 1749.558289 | |
| 65-74 | 0.000182273 | 0.0008196 | 0.00090846 | 4336.098128 | 2705.4794 | 2363.18426 | |
| 75-84 | 0.00058816 | 0.00185972 | 0.00184476 | 7332.634523 | 2699.4567 | 2245.240107 | |
| 85+ | 0.001553469 | 0.00286603 | 0.00275206 | 8161.683723 | 1553.8896 | 1315.579881 | |
| | | | | 22156.82977 | 9529.5546 | 9176.85883 | 40863.243 |
| | | | | | | Deaths expected | **43,187** |

US Population: 328,239,523

Population NH white, black 306,867,627 Remaining pop: 21,371,896

Ratio: 1.06964533

Sources:

German mortality Robert Koch Institut - COVID-19. https://www.rki.de/EN/Content/infections/epidemiology/outbreaks/COVID-19/COVID19.html. Accessed September 29, 2020.

US mortality by race and ag Deaths involving coronavirus disease 2019 (COVID-19) by race and Hispanic origin group and age, by state | Data | Centers for Disease Control and Prevention. https://data.cdc.gov/NCHS/Deaths-involving-coronavirus-disease-2019-COVID-19/ks3g-spdg. Accessed September 15, 2020
